# A scalable invariant causal discovery framework reveals direct genetic and clinical determinants of complex human traits

**DOI:** 10.1101/2024.08.29.24312763

**Authors:** Liangying Yin, Menghui Liu, Yujia Shi, Jinghong Qiu, Hon-Cheong So

## Abstract

Identifying genetic and clinical factors that exert *direct causal* effects on complex traits is central to precision medicine, yet remains a major challenge. Association-based methods cannot distinguish cause from consequence, and existing causal frameworks are limited by stringent assumptions or computational intractability in high-dimensional settings. Invariant causal prediction (ICP) offers a principled alternative but has seen minimal application in genetic epidemiology due to exponential complexity.

Here, we introduce the Invariant Generalized Covariance Measure (I-GCM), a scalable framework that integrates ICP principles with machine learning to identify direct causal variables. I-GCM combines PC-simple screening with a generalized covariance measure (GCM) computed via gradient-boosted trees to test invariance across environments, capturing both linear and nonlinear dependencies. In simulations (up to 3,000 variables, 50,000 samples), I-GCM achieved PPV of ∼0.84–1.00 and specificity ≥0.99, outperforming PC-simple by 0.21–0.29 in precision with comparable sensitivity. Applying I-GCM to the UK Biobank, we identified direct causal genes and clinical risk factors for COVID-19 susceptibility and severity and for HDL and triglyceride levels, with many findings supported by existing literature and pathway enrichment. Notably, *JAK1* was identified as a direct causal gene for severe COVID-19, consistent with clinical use of the JAK1-targeting therapy baricitinib.

To our knowledge, this is among the first applications of invariance-based causal discovery to genetic epidemiology at a biobank scale. By bridging statistical causality and translational medicine, I-GCM offers a scalable, interpretable, and high-precision framework for mechanistic discovery and therapeutic target prioritization.

## Introduction

Determining which genetic and clinical factors exert direct causal effects on complex traits is central to precision medicine and to understanding disease mechanisms, yet remains a major unsolved challenge. Statistical and machine learning approaches have transformed biomedical data analysis by uncovering associations, but such associations alone cannot distinguish causes from consequences. Identifying *direct causal* factors is crucial as they serve as reliable biomarkers or therapeutic targets, compared to associations. Despite the rapid growth of large-scale genomic and phenotypic datasets, methods that can robustly identify these factors in realistic, heterogeneous data remain scarce. Causal inference offers a promising path toward disentangling cause from correlation, and has gained momentum across many scientific fields^1–5^. However, practical frameworks that can reveal direct causal variables in complex biomedical contexts are still limited^6^. Consequently, the field still lacks scalable, interpretable methods that can uncover true causal determinants directly from large, heterogeneous biomedical datasets.

A key challenge lies in identifying the true ***direct causal*** variables of an outcome from large-scale, heterogeneous observational data. In earlier work, Bühlmann et al.^7^ presented a method (PC-simple), which utilized partial correlation screening, to infer the linear causal relations between covariates and the target from observational data. However, it was not designed to incorporate multiple data sources for causal inference. Also, for PC-simple, assumption of a linear model is typically required. A previous work by Peters et al.^8^ proposed to exploit the invariance property of a causal linear model (ICP) from different experimental settings/environments (i.e., observational and interventional data) to uncover the direct causal variables for a target. Heinze-Deml et al.^9^ later extended this framework to accommodate nonlinear causal relations (nonlinear ICP). While these approaches provide theoretical insight, their computational complexity increases exponentially with variable dimension (2^*q*^) and their statistical power declines when the true causal set size is large. These constraints make them infeasible for high-dimensional biomedical dataset, leaving a critical gap in our ability to perform robust causal discovery in modern biobanks. Addressing these challenges is critical as modern biobank-scale datasets increasingly demand causal frameworks that can handle both complexity and scale.

Despite their theoretical advantages, ICP and related causal frameworks remain very rarely applied in biomedical, genomic (e.g., GWAS), and clinical research. Existing attempts to infer causality in clinical and molecular data are often based on instrumental variable frameworks, such as Mendelian Randomization (MR) and summary-data-based MR (SMR) utilizing eQTLs, or traditional observational adjustments like multivariate regression and propensity score matching. However, these conventional frameworks present their own limitations. MR-based approaches rely on stringent instrumental variable assumptions (e.g., absence of horizontal pleiotropy) and typically assess targeted pairwise causal relationships rather than discovering (possibly multiple) direct causal parents from high-dimensional features. On the other hand, multivariate adjustment and propensity score matching control for measured confounding to estimate conditional associations, which do not necessarily identify *direct* causal variables. Distinguishing direct causal parents from indirect or downstream mediators is of critical scientific and clinical importance; for example, a gene that exerts a direct causal effect on a disease outcome usually represents a far more promising candidate for drug target discovery than one whose influence is indirect or mediated through secondary pathways.

Here, we bridge the above research gaps with Invariant Generalized Covariance Measure (I-GCM), a scalable causal discovery framework that integrates invariance-based causal inference with flexible machine learning models to identify direct causal variables in high-dimensional biomedical data. While ICP provides a theoretically sound foundation for causal inference, existing implementations are largely limited to low-dimensional applications and have seen minimal use in biomedical and clinical research, where datasets are often high-dimensional and heterogeneous.

I-GCM extends the applicability of ICP by incorporating a scalable variable selection step followed by a robust causal inference procedure. In the first stage, the PC-simple algorithm is employed to reduce dimensionality and retain variables likely to include direct causal factors. In the second stage, we introduce a generalized covariance measure (GCM), computed using gradient-boosted trees, to test for invariance of causal associations across multiple environments or data sources. This approach enables detection of both linear and non-linear dependencies, enhancing flexibility and robustness. Importantly, by exploiting heterogeneity across environments, I-GCM aims to substantially reduce false positive rates, an essential consideration in biomedical settings where experimental validation is usually resource-intensive.

To demonstrate the utility of I-GCM, we conducted extensive simulations and applied the method to the UK Biobank, a large-scale resource integrating genetic, clinical, and environmental data. We identified direct genetic and clinical risk factors for several complex traits and diseases, including COVID-19 susceptibility and severity, as well as lipid traits such as high-density lipoprotein (HDL) and triglyceride (TG) levels. These findings highlight the method’s capacity to yield biologically and clinically meaningful insights.

Our primary contributions can be summarized as follows:

1. **Scalable methodology for high-dimensional causal discovery** We combine machine learning approaches with the principles of ICP to identify direct causal variables in the presence of a large number of covariates. Previous ICP applications or methods primarily focus on the case when only a limited of covariates are present; I-GCM is specifically designed to scale to the high-dimensional datasets (such as omics and clinical data) that are common in modern biomedical studies.
2. **Novel application to genetic epidemiology** While ICP is a theoretically sound and powerful framework, it has seen minimal substantive applications in large-scale clinical studies, and we are unaware of any previous applications in genetic epidemiology. Here, we bridge this gap by applying I-GCM to the UK Biobank to successfully identify direct causal genes and clinical risk factors underlying complex traits/disorders.
3. **Translational and biological insights** We identify specific genetic and clinical factors that exert direct causal effects on COVID-19 susceptibility and severity, as well as lipid traits including high-density lipoprotein (HDL) and triglyceride (TG) levels. These findings yield biologically and clinically meaningful insights with the potential to inform risk prediction and therapeutic target prioritization. As an example, our framework pinpoints *JAK1* as a direct causal gene for severe COVID-19. Notably, *JAK1* is the target of baricitinib, an established clinical treatment for COVID-19. As such, this finding serves as a compelling proof-of-concept for our method’s translational utility.

To our knowledge, this is among the first applications of invariance-based causal discovery to human genetic-epidemiological and biobank-scale data. Our study demonstrates that the integration of invariant principles with machine learning not only improves the scalability and accuracy of causal discovery in high-dimensional settings, but also enables the identification of stable, generalizable causal relationships with direct implications for translational medicine.

## Methods

### I-GCM design

I-GCM comprises two steps: variable selection, and direct causal variable set identification (Fig.1). For variable selection, we proposed to employ the PC-simple^7^ algorithm, which utilizes ordered independence test to screen potential causal variables for the outcome under study. Following this, we employ our proposed metric, i.e., invariant generalized covariance measure, to infer the ultimate causal variable set for our studied trait from the preselected variables. This metric leverages the invariance property of causal relations across different experimental settings to identify direct causal variables. In the following sections, we will delve into the details of this method.

**Fig. 1.**
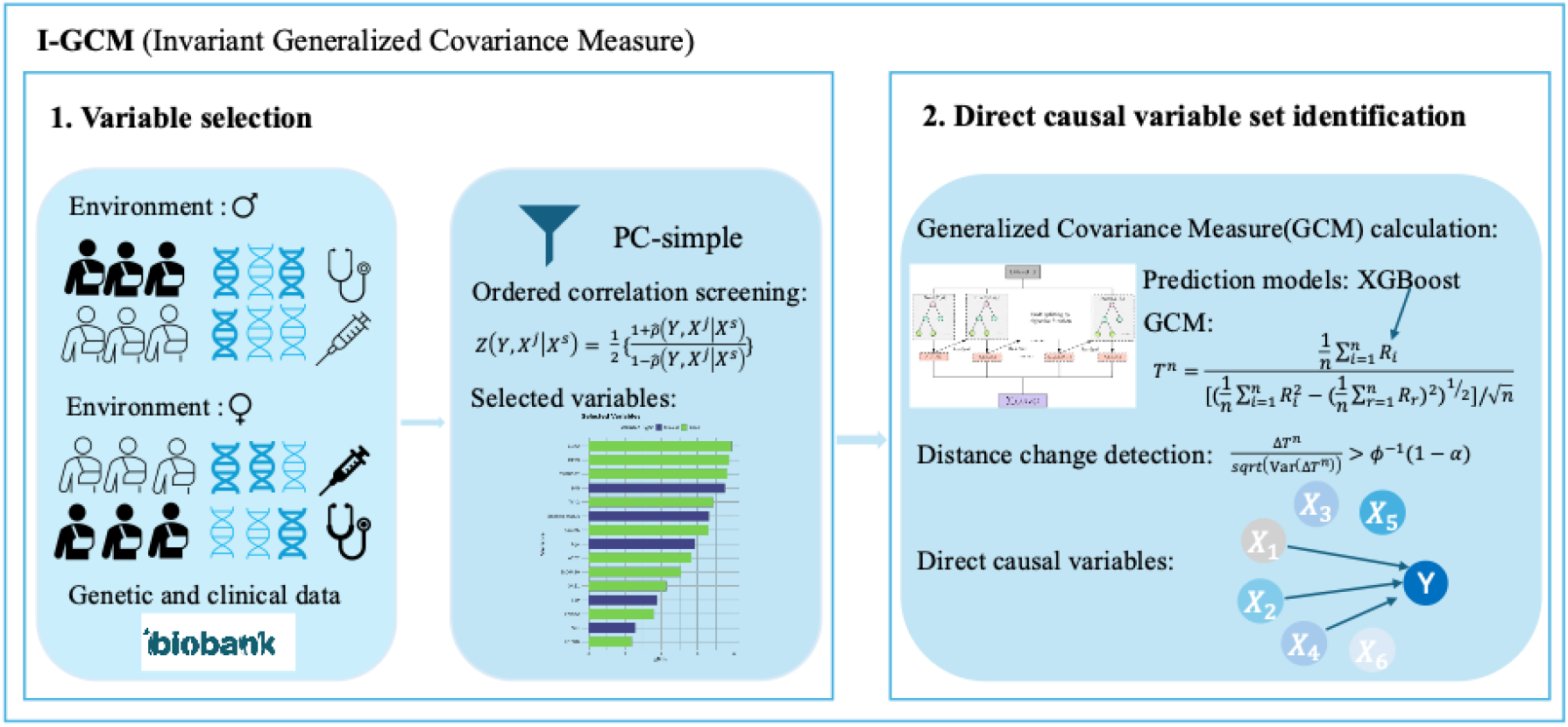
Schematic illustration of the I-GCM algorithm. I-GCM consists of two stages: (1) variable selection and (2) direct causal variable set identification. 1. In the first stage, we apply the PC-simple algorithm to pre-select candidate variables that are potentially direct causes of the outcome. This step reduces the dimensionality of the problem and narrows the search space for subsequent causal inference. 2. In the second stage, we adopt a backward feature selection strategy to iteratively evaluate nested candidate sets. At each iteration, we compute the generalized covariance measure (GCM) for two consecutive candidate variable sets using the residuals from XGBoost prediction models. We then assess whether the distance change in GCM between the candidate sets. Is statistically significant. By leveraging the invariance property of causal relations across different experimental settings, the algorithm can identify the ultimate direct causal variable set.

### Variable selection via PC-simple

As mentioned earlier, we utilized the PC-simple algorithm to first perform variable selection. It employs partial correlation screening to eliminate irrelevant variables for the target variable. We first review the original PC-simple algorithm. Let *X* = [*X*^1^,*X*^2^..*X*^p^] be a *n* ×*p* matrix for *n* observations with *p* variables, *Y* be a vector for *n* observations. Suppose is defined by the following generative model:

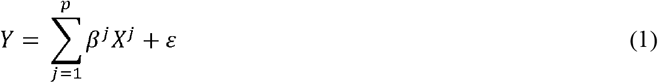

Here, ε indicates the noise item that is independent of *X*^*j*^ and follows a multivariate normal distribution (∈ ∼*N*(0, Σ)). Variable with a non-zero β^*j*^ is the true direct causal variable for *X*^*j*^. If is independent of β^*j*^ =0, i.e.,, then we have:

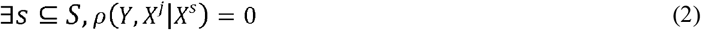

Here, *S* = {1,2,…,*j* + 1,…,*p*}defines the set including variables excluding *X*^*j*^, is a subset of *S*, ρ(Y, *X*^*j*^ |*X*^*S*^) denotes the partial correlation between and. We then perform Fisher’s Z-transform:

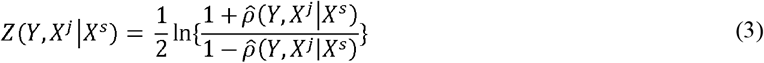

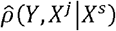 indicates the estimated partial correlation from data. The null hypothesis of independence would be rejected if

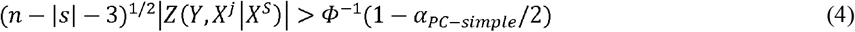

*n* denotes the sample size, |*S* | is the cardinality of the set, Φ ^−1^denotes the inverse of the standard normal cumulative distribution function. Recursively performing partial correlation screening with increased order could exclude irrelevant variables from previous candidate variables set until they do not vary anymore.

In this study, we set the maximum order of |*s* | to 3 and α_*PC*−*simple*_ to 0.05 for the feature screening process. This implies that variables which survive the 3-order partial correlation screening will be retained for further analysis. For more details about this method, please refer to ^7^. The feature dimension *q* will be dramatically reduced after employing the PC-simple algorithm for feature selection.

### Causal variable set identification

While the PC-simple algorithm outperforms commonly used feature selection methods (e.g., Lasso, elastic net, etc.) in selecting causally relevant variables for biomedical data, it still struggles with high false positive rates in high-dimensional data and non-linear relations. To address these challenges, we propose a novel framework that leverages the invariance property of causal models to identify a direct causal variable set. Causal relations are universal and robust across different experimental settings or environments. The key idea underlying ICP is that the conditional distribution of the target variable *Y* given its direct causes (*Xs*) remains invariant, if we intervene on other variables in the model, except the target itself. Put it in another way, given the complete causal set, the conditional distributions for the target variable across various experimental settings/environments should be identical. This also implies predictions from a causal model will remain consistent across different environments.

In biomedical studies, ‘omics’ measurements are often made under a combination of interventional and observational settings, or under different interventions, for example knockout of different genes, different drug treatments etc. The ICP approach is a natural choice for causal inference in such scenarios, but it is also useful in observational settings with distinct ‘environments’. Intuitively, the causal structure or components should be consistent across different sub-populations, while non-causal components may vary.

In a previous work, Shah and Peters^10^ proposed to use generalized covariance measure (GCM) to detect the causal relationships between variables. We will extend this concept to reliably identify causal variable set in high-dimensional data settings.

#### Generalized covariance measure (GCM)-based causal variable identification

For a given distribution of random variables (*X* ,*Y* ,*Z*) we can always decompose the distribution into the following equations:

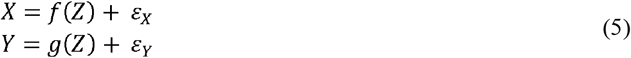

Notably, *Z* can either be a single variable or a set of variables. We employed XGBoost (gradient boosted trees)^11^ to build the prediction models *f* (*Z*) and *g* (*Z*). Note that there is no restriction on the type of regression or machine learning models for *f* (*Z*) and *g* (*Z*). In brief, XGBoost is a supervised learning method attempting topredict the target by combining the estimates from a sequential of simpler and weaker tree models. Each new tree is trained based on the residuals from the previous tree using gradient descent to minimize the loss. Given *n* observations for the variables *(x*_*i*_,*y*_*i*_,*z*_*i*_) (*i* = 1,2,…*n*), we could calculate the product between residuals(*R*) from the prediction functions for each observation, i.e.,

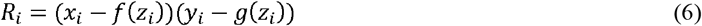

The normalized sum of *R*_*i*_ (*T*^*n*^)(a.k.a., generalized covariance measure (GCM)) could be represented as follows:

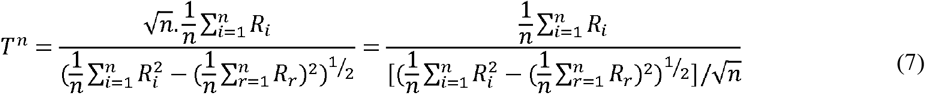

*T*^*n*^ could be utilized to test the null hypothesis that *X* and *Y* are conditional independent given variable(s) Z. Shah and Peters^10^ proved that *T*^*n*^ is asymptotically standard normal. The null hypothesis would be rejected if | *T*^*n*^| has a larger value, i.e.,

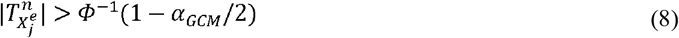

The parameter α_*GCM*_ is set to a default value of 0.05.

This method can be used to detect whether a single variable is causally relevant to the target variable. If the conditional set *Z* encompasses all direct cause for the target variable, then *X* and *Y* are conditionally independent given variable Z. In our case, *X* will be replaced by the environmental variable *E*, i.e., we will test if the target *Y* is independent of the environment *E*, given a set of covariates *Z*^9^.

If *Z* contains all direct causal variables, *Y* should be invariant across or independent of *E*. It is worth noting that even if *Z* includes some irrelevant variables, the above-mentioned null hypothesis remains valid. Direct application of this method for detecting reliable causal variable set may result in the identification of a set with superfluous irrelevant variables. To mitigate this, we proposed to extend this concept and combine it with the invariance property of causal relations to identify reliable causal variables set for the target.

As discussed above, for a given environment variable *E*, the computed GCM between *E* and the target should be constant or similarly close to zero when conditioned on the full set of direct causes, even if some other (non-directly causal) variables are included. Throughout, the environment variable enters as one argument of a single conditional-independence test: for each candidate set *S*_*j*_ we test *E* ⊥ *Y*|*S*_*j*_ on the full sample, using the per-subject residual products of *E* on *S*_*j*_ and *Y* of on *S*_*j*_ (equations 6–7). Continuous environment variables are dichotomized at their median before entering the test; discrete ones are used as observed. However, if we exclude some direct causes from the conditional set, the calculated GCM between *E* and the target is expected to divert away from the null. In other words, significant change of the calculated GCM would be observed between the reduced conditional set and full conditional set. If we exclude some irrelevant variables in the conditional set, the calculated GCM should remain stable. Notably, a direct causal variable for the target can also act as an environment variable^8^. If this is the case, the aforementioned statements still hold; under this scenario, the computed GCM between *E* and the target will be non-zero, but the relationship should be constant, if conditioned on other direct causes. The “full set of direct causes” does not include the environment variable in this case.

We employed backward feature selection to sequentially drop variables in the conditional set and calculate the GCM for each conditional set. The variables were ordered by the derived *Z*_*min*_ value from the PC-simple algorithm in descending order (i.e. the *least* likely causal variables are ranked last, and will be removed from the set first) before performing the backward feature selection. Here *Z*_*min*_ refers to the minimum *z-*statistic in the series of conditional tests from PC-simple; a higher *Z*_*min*_ in general reflects stronger causal relationships. Here we propose a backward elimination procedure as it is practically impossible to enumerate all combinations of all covariates as direct causal variables. This may be considered a ‘greedy’ approach, but it can greatly reduce the computational burden to make modeling of high-dimensional data possible.

The change in the distance of GCM from zero between two consecutive conditional sets (Δ*T*^*n*^) could be represented as follows:

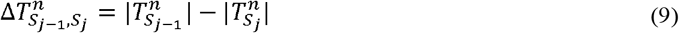

Here we used the absolute value as we are mainly concerned about the distance of GCM from zero, which reflects the degree of independence of *E* and *Y* (GCM=0 indicates complete independence), conditioned on other estimated direct causes. The variance of the distance change of GCM 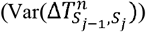 under the null can be estimated from the following:

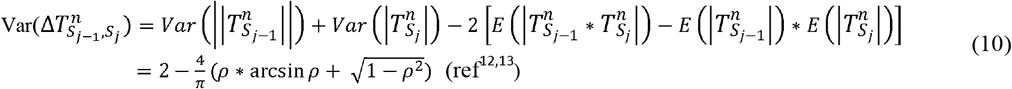

Where 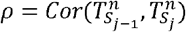 indicates the correlation between product of residuals calculated from conditional set *S*_*j −* 1_ and *S*_*j*_, *S*_*j −* 1_ and *S*_*j*_ respectively denote two consecutive conditional set with *j* − 1 and *j* variables. Under the null hypothesis that the j-th variable is not a direct cause, 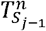 and 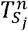 are asymptotically bivariate normal with unit variances and a common mean μ, so that 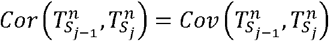. Equation 10 takes μ = 0, which holds when the environment variable E is not itself a direct cause of Y^12,13^. When E is a direct cause of Y, μ ≠ 0 and the exact variance of ΔT is its non-central counterpart (Supplementary Text). We use equation 10 throughout; its calibration when *E* is a direct cause, and the comparison with the non-central form, are assessed by simulation (see Simulation results). After replacing both 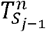 and 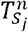 with equation 7, we have:

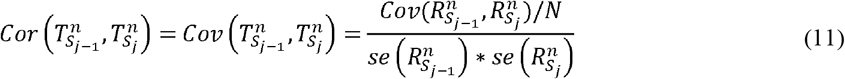

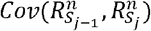 indicates the covariance between product of residuals calculated from conditional set *S*_*j*−1_ and *S*_*j*_,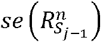 and 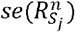 respectively indicate the standard error for product of residuals calculated from conditional set *S*_*j*−1_ and *S*_*j*_, *N* and is the sample size of the target dataset. For more details, please refer to the supplementary text. The null hypothesis of no significant distance change in GCM between two consecutive conditional sets would be rejected if:

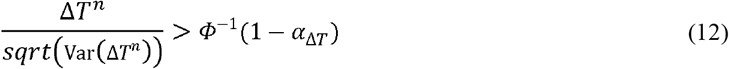

Here Φ ^−1^ denotes the inverse of the standard normal cumulative distribution function; the test is one-sided because removing a direct cause from the conditional set is expected to increase the distance of the GCM from zero. It should be noted that the above does not account for multiple testing, as typically repeated hypothesis tests need to be performed. We recommend a principled approach for formal statistical inference by stability selection, as described below.

Algorithm 1 summarizes how we may identify a reliable direct causal variable set by I-GCM. Notably, *α* _Δ*T*_ and *T* may vary between different data settings, where *T* refers to the threshold for Δ*T*^*n*^. For low-dimensional settings, we may simply set *α* _Δ*T*_ to 0.05 and *T* to 0. A more stringent criteria is recommended for high-dimensional settings. In this study we used a tiered rule keyed to the screened set size q: (*α* _Δ*T*_, T) = (0.05, 0) for q ≤ 60; (0.01, 0) for 60 < q ≤ 110; (0.005, 0.1) for 110 < q ≤ 160; (0.005, 0.2) for 160 < q < 210; and (0.001, 0.3) for q ≥ 210. The selected set is the largest candidate set for which both criteria hold; if no step satisfies them, the full screened set is returned. Note that if the investigator wishes to experimentally follow up a limited number of genes, the thresholds may be adjusted such that the number of genes left matches with the number planned for further follow-up studies. As detailed below, a formal statistical approach (stability selection) can be employed to control the expected number of false positive findings.

##### Algorithm 1: I-GCM for causal variable set identification

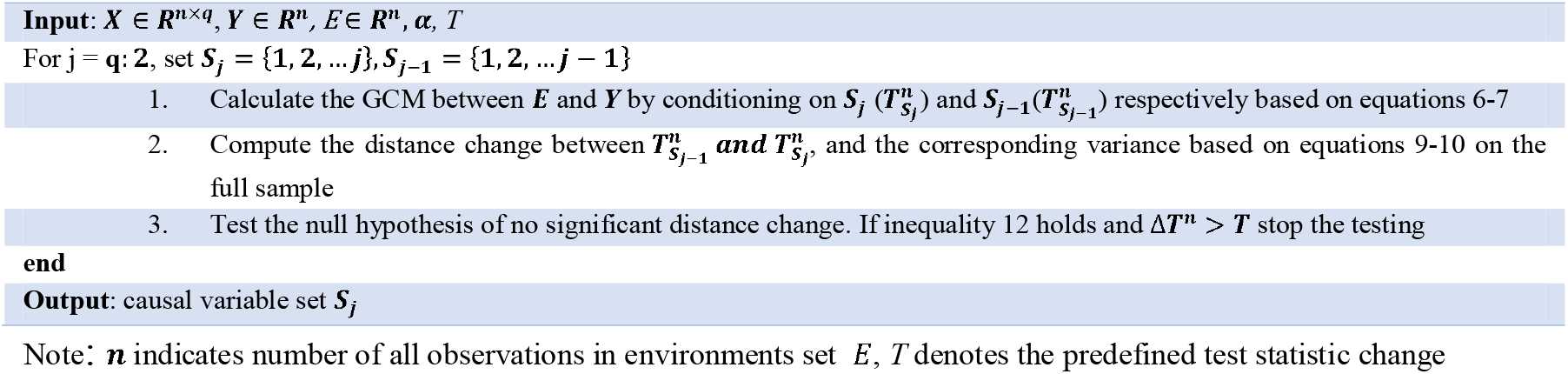

#### Stability selection for statistical inference

We propose the stability selection method ^14^ as a principled approach to formally control the expected number of falsely selected variables. In brief, this approach involves repeated subsampling of the data, and variables that are consistently selected across subsamples are more likely to be true positives. Meinshausen and Bühlmann^14^ established a formula to control for *E*(*V*), the expected number of false positives.

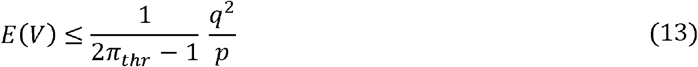

*π*_*thr*_ defines the selection probability cutoff (selection probability refers to the proportion of times a certain feature is selected across all sub-samples),*q* denotes the average number of identified variables, while *p* indicates the original number of variables. If stability selection is applied after PC-simple pre-screening, this bound should be regarded as heuristic: q is counted among the screened variables whereas p refers to the pre-screening count, and the selection events are not those for which the bound was derived.

In addition, stability selection can help identify the optimal parameters (*α* _Δ*T*_ and *T*) for analysis, as different parameters will result in different *E*(*V*). The researcher may adjust *α* and *T* accordingly to achieve a desired level of *E*(*V*). Alternatively, the *E(V)* can be controlled given a combination of parameters (*α* _Δ*T*_ and *T*). In this study we used the tiered rule described above, keyed to the screened set size q (q > 160 in our data applications).

We conducted a further pilot study to illustrate the application and usefulness of stability selection, under a moderate data dimension: *p*=1000 ,*n* =50000 We set the iteration number to 50, subsampling *80%* of the observations for each iteration. We applied a selection probability threshold of 0.9 to determine which variables were consistently selected across iterations. As suggested by Meinshausen et al., *π*_*thr*_ has only minor effect on the results when it falls into a reasonable range [0.6, 0.9]

### Simulation

To verify the validity of our proposed framework, we simulate different scenarios with varying total variable number (*p*), sample sizes (*n*), environment variable types (*t*_*e*_). Specifically, for each scenario, we randomly generated a directed acyclic graph (DAG) with the nodes representing the variables and the causal effects randomly generated from a given range following a uniform distribution. We employed the function “rmvDAG” in the R package “pcalg” ^15^ to realize this. The original function is designed to generate multivariate data with dependency structures specified by a given DAG. To make it adaptive to binary variables, we employed a predefined liability threshold (i.e., 0.3) to convert the simulated continuous variable into a binary one. As the threshold was solely used for variable type transformation, it does not affect the validity of I-GCM. The number of direct causes (*p*_*c*_) in each scenario was determined by graph density. To align with real-world application scenarios, we set the graph density to 0.05. We considered the following combination of settings:

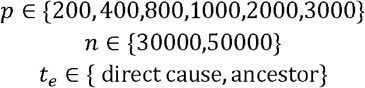

We generated data from randomly chosen DAG following linear Gaussian structural equation models. Also, we generated scenarios with a mixture of linear and non-linear relations with *p*∈{2000,3000}. For scenarios with non-linear relations, we randomly selected ∼2.5% of the overall direct causal variables for the target variable and replaced the linear relations with quadratic forms. The simulated datasets were then utilized to identify the causal variable set for the chosen target. For all simulated scenarios, the last variable was chosen as the target variable. The environment variable was randomly selected from the direct causes or ancestors of the target.

We evaluated the efficacy of I-GCM in identifying the direct causal variables for the target variable. We utilized 4 different metrics to evaluate the performance, i.e., positive predictive value (PPV, a.k.a., precision), negative predictive value (NPV), sensitivity, specificity. PPV represents the proportion of identified direct causal variables that are true ones while NPV denoted the same for non-direct causal variables. Sensitivity measures the ability in detecting the true direct causal variables while specificity gauges the ability to identify actual non-direct causal variables. We additionally report F1 and F0.5 scores.

In addition, we compared the performance of I-GCM with the PC-simple algorithm in terms of recovering the direct causes for the target. For I-GCM, the environment variable was used directly in the GCM test as described above. Besides, we evaluated the performance of our proposed under unobserved confounding. Specifically, we extended previous simulation framework by adding latent variable that affects both a subset and the target outcome *Y*. Furthermore, we benchmarked our causal inference method against traditional association-based method, i.e., Lasso combined with stability selection. To further quantify variability across problem instances, all scenarios were repeated over 10 independent replicates, with the DAG, the simulated dataset and the environment node redrawn under fresh random seeds in each replicate. Performance metrics are reported as the average across replicates.

### Real data application

We applied I-GCM to the UK-Biobank(UKBB) dataset, which contains GWAS data and clinical variables, to identify (direct) causal variable set for different phenotypes, i.e., COVID-19 infection, severe COVID-19, high density lipoprotein (HDL) and triglycerides (TG). Analyses were restricted to UK Biobank participants with a recorded SARS-CoV-2 test result (n = 154,749); untested participants were excluded. For severe COVID-19, cases were test-positive participants who were hospitalised or who died (ve-pos-inpatient and ve-pos-death), and controls were test-negative participants together with test-positive participants managed as outpatients. For COVID-19 infection, cases were all test-positive participants and controls were test-negative participants. Because the study population is conditioned on having been tested, and because the severe-COVID control group combines uninfected and non-hospitalised infected individuals, these analyses are subject to test-based selection and the resulting estimands should be interpreted accordingly. Analyses were not restricted by ancestry; ancestry principal components were not included among the conditioning variables. Self-reported ethnicity was retained as a candidate covariate but is interpreted as a social and contextual marker, capturing differences in exposure, occupation, healthcare and testing access, and genetic ancestry rather than a biological cause. The expression models are autosome-only, so X-linked sex differences do not affect the sex-environment analyses.

All the clinical variables included were extracted from the UK-Biobank dataset under application no. 37268. For the lipid traits, several candidate variables are measured from the same blood sample as the outcome and are compositionally or metabolically coupled to it. Total cholesterol mathematically contains HDL-C (TC = HDL-C + LDL-C + VLDL-C); apolipoprotein A is the principal structural protein of the HDL particle; and apolipoprotein B, LDL, lipoprotein(a) and triglycerides are linked to the outcome through shared lipoprotein metabolism. These variables were retained in the candidate set, but any selection among them is interpreted as a compositional or metabolic association rather than as a direct causal effect. Causal interpretation of the lipid analyses is therefore restricted to the non-lipid clinical variables and to the identified genes.

Our input data comprises both clinical variables and imputed gene expression data. In this study, we employed “PrediXcan”^16^ to impute tissue-specific gene expression levels from the genotypic data of the UK-Biobank subjects. In brief, PrediXcan was based on a prediction model for gene expression levels using elastic-net based regression model on GTEx (a reference dataset with available genotype and gene expression). The genotypic data of the UK-Biobank subjects were then used to “impute” the tissue-specific gene expression levels. The continuous clinical covariates were directly extracted from the UK-Biobank. The binary covariates were defined by ICD10-coded disease in the UK-Biobank. We incorporated imputed gene expression levels from whole blood and the lung for the analysis. Table S1 summarizes the studied phenotypes. For all application scenarios using UK Biobank, we chose sex as the environment variable. This choice is predicated on the well-established physiological and metabolic differences between males and females^17,18^, which can create distinct biological contexts. We hypothesized that while these sex-specific environments might alter the distribution of covariates or the apparent effects of non-causal variables, the conditional distribution of the outcome given its true direct causal variables should remain invariant across sexes, as per the principles underlying ICP. This approach allows us to leverage naturally occurring heterogeneity within the population to refine the identification of direct causal factors.

To further validate the reliability of I-GCM in identifying (direct) causal variables, we compared its performance with the PC-simple algorithm in identifying potential targets for COVID-19. Specifically, we assessed whether the gene sets identified by our method had an equivalent chance of being listed as targets in Open Targets. The targets associated with COVID-19 were downloaded from Open Targets Platform, which provides measures of relevance (ranging between 0 and 1) between potential targets and COVID-19 based on various factors such as associations with known drugs, hits in relevant GWAS, etc.

To gain a deeper understanding of the biological mechanism underlying the identified causal genes for the target outcome, we performed pathway enrichment on the identified gene set. More specifically, an over-representation analysis was conducted on the identified causal genes using the web-based tool “ConsensuspathDB” ^19,20^. Furthermore, drug enrichment analyses were carried out to identify drugs related to COVID-19 infection and severe COVID-19 using the CREEDS drug-signature library; enrichment p-values were adjusted for multiple testing across all tested compounds.

## Results

### Overview

We proposed a novel causal inference framework, invariant generalized covariance measure(I-GCM), which integrates flexible machine learning method with the principle of invariant causal prediction to identify direct causal variables for a given outcome variable in high-dimensional settings (Fig.1). We first evaluated I-GCM through extensive simulations, where it yielded high precision (PPV > 0.90 in most simulated scenarios) and specificity while maintaining comparable sensitivity. Its PPV advantage over PC-simple widened as the number of candidate variables increased. We then applied I-GCM to the UK-Biobank, analyzing genetic and clinical data to identify causal factors for COVID-19 infection/severity and lipid traits (HDL, Triglycerides). The analysis successfully recovered many known risk factors, validating the method’s real-world performance, and revealed additional putative causal genes and biological pathways supported by existing literature.

### Simulation results

Table 1 summarizes the simulation results across scenarios, and Table 2 reports the corresponding evaluation metrics; both report a single draw per scenario. I-GCM achieved PPV ≥ 0.84 and specificity ≥ 0.99 in every scenario (Table 2) while retaining satisfactory sensitivity (Fig. 2a). Performance remained stable as the number of input variables increased, and F1 and F0.5, which weigh both PPV and sensitivity, were generally higher for I-GCM than for PC-simple. As expected, sensitivity improved with larger sample sizes.

**Table 1.** Simulation results for our proposed I-GCM and the PC-simple algorithm.

| **Overall sample size** | **Environment variable type** | **No. of input variables** | **No. of overall true causal** | **PC-simple** |  |  | **I-GCM** |  |  |
|---|---|---|---|---|---|---|---|---|---|
|  |  |  |  | total no. | true causal | false causal | total no. | true causal | false causal |
| 50000 | Direct cause | 200 | 11 | 13 | 9 | 4 | 7 | 7 | 0 |
| 50000 | Direct cause | 400 | 21 | 27 | 17 | 10 | 15 | 15 | 0 |
| 50000 | Direct cause | 800 | 39 | 38 | 28 | 10 | 31 | 28 | 3 |
| 50000 | Direct cause | 1000 | 51 | 57 | 43 | 14 | 46 | 42 | 4 |
| 50000 | Ancestor | 1000 | 51 | 59 | 46 | 13 | 48 | 45 | 3 |
| 30000 | Direct cause | 1000 | 51 | 54 | 36 | 18 | 33 | 33 | 0 |
| 50000 | Direct cause | 2000 | 114 | 121 | 87 | 34 | 90 | 82 | 8 |
| 50000 | Ancestor | 2000 | 114 | 110 | 87 | 23 | 99 | 86 | 13 |
| 30000 | Direct cause | 2000 | 114 | 110 | 77 | 33 | 64 | 64 | 0 |
| 50000 | Direct cause | 3000 | 163 | 215 | 122 | 93 | 111 | 108 | 3 |
| 50000 | Ancestor | 3000 | 163 | 216 | 121 | 95 | 128 | 110 | 18 |
| 30000 | Direct cause | 3000 | 163 | 168 | 114 | 54 | 115 | 97 | 18 |

**Table 2.** Comparison of different evaluation metrics between our proposed I-GCM and the PC-simple algorithm.

| Overall sample size | Environment variable type | No. of input variables | No. of overall true causal | PC-simple |  |  |  |  |  | I-GCM |  |  |  |  |  |
|---|---|---|---|---|---|---|---|---|---|---|---|---|---|---|---|
|  |  |  |  | PPV | Sensitivity | NPV | Specificity | F1-score | F0.5-score | PPV | Sensitivity | NPV | Specificity | F1-score | F0.5-score |
| 50000 | Direct cause | 200 | 11 | 0.692 | 0.818 | 0.989 | 0.979 | 0.750 | 0.714 | 1.000 | 0.636 | 0.979 | 1.000 | 0.778 | 0.897 |
| 50000 | Direct cause | 400 | 21 | 0.630 | 0.810 | 0.989 | 0.974 | 0.709 | 0.659 | 1.000 | 0.714 | 0.985 | 1.000 | 0.833 | 0.926 |
| 50000 | Direct cause | 800 | 39 | 0.737 | 0.718 | 0.986 | 0.987 | 0.727 | 0.733 | 0.903 | 0.718 | 0.986 | 0.996 | 0.800 | 0.859 |
| 50000 | Direct cause | 1000 | 51 | 0.754 | 0.843 | 0.990 | 0.987 | 0.796 | 0.770 | 0.913 | 0.824 | 0.990 | 0.998 | 0.866 | 0.894 |
| 50000 | Ancestor | 1000 | 51 | 0.780 | 0.902 | 0.995 | 0.986 | 0.837 | 0.802 | 0.938 | 0.882 | 0.994 | 0.997 | 0.909 | 0.926 |
| 30000 | Direct cause | 1000 | 51 | 0.667 | 0.706 | 0.984 | 0.981 | 0.686 | 0.674 | 1.000 | 0.647 | 0.981 | 1.000 | 0.786 | 0.902 |
| 50000 | Direct cause | 2000 | 114 | 0.719 | 0.763 | 0.988 | 0.980 | 0.740 | 0.727 | 0.911 | 0.719 | 0.983 | 0.999 | 0.804 | 0.865 |
| 50000 | Ancestor | 2000 | 114 | 0.791 | 0.763 | 0.986 | 0.988 | 0.777 | 0.785 | 0.869 | 0.754 | 0.985 | 0.993 | 0.807 | 0.843 |
| 30000 | Direct cause | 2000 | 114 | 0.700 | 0.675 | 0.980 | 0.983 | 0.687 | 0.695 | 1.000 | 0.561 | 0.974 | 1.000 | 0.719 | 0.865 |
| 50000 | Direct cause | 3000 | 163 | 0.567 | 0.748 | 0.985 | 0.967 | 0.645 | 0.596 | 0.973 | 0.663 | 0.981 | 0.999 | 0.789 | 0.890 |
| 50000 | Ancestor | 3000 | 163 | 0.560 | 0.742 | 0.985 | 0.967 | 0.638 | 0.589 | 0.859 | 0.675 | 0.982 | 0.994 | 0.756 | 0.815 |
| 30000 | Direct cause | 3000 | 163 | 0.679 | 0.699 | 0.983 | 0.981 | 0.689 | 0.683 | 0.843 | 0.595 | 0.977 | 0.994 | 0.698 | 0.778 |

**Fig. 2.**
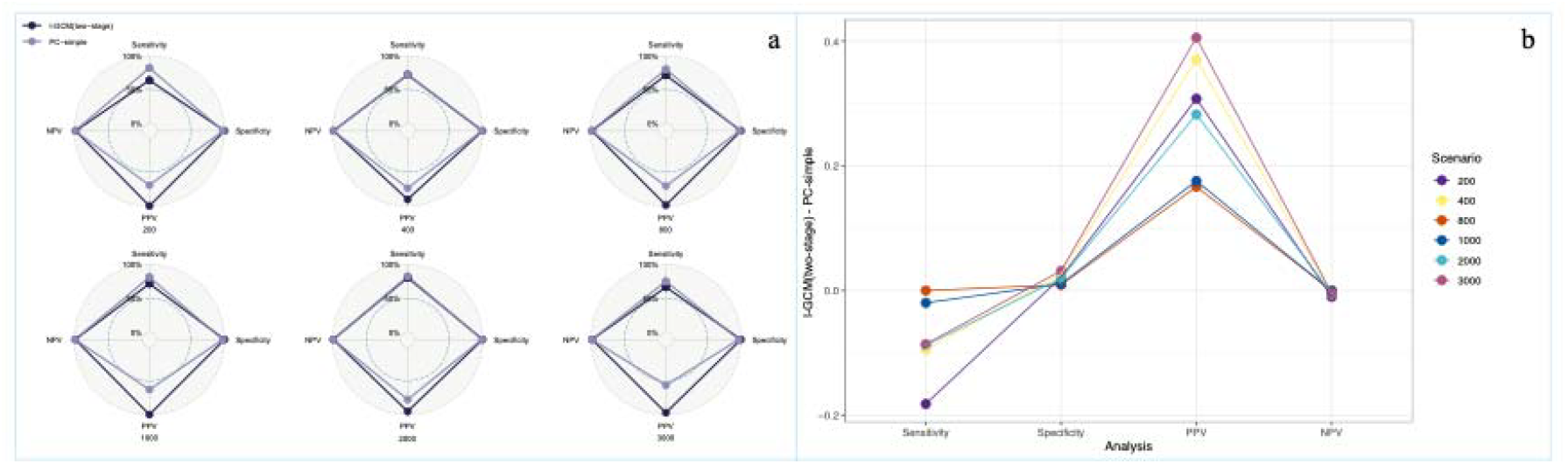
Performance comparison between our proposed I-GCM and the PC-simple algorithm in causal variables discovery. a. Overall performance of the proposed I-GCM method compared to the PC-simple algorithm across simulated scenarios. b. Differences between I-GCM and PC-simple across 4 evaluation metrics

Both direct cause and ancestor variables were selected as environment variables to identify direct causal variables in our simulations. They demonstrated comparable performance in terms of the prediction metrics (Table 2). The direct cause seemed to be a more potent candidate for environment variable in eliminating false positives. We hypothesize that interventions on direct cause probably induced more discernable and detectable heterogeneity in the corresponding subsets than ancestors, aiding in distinguishing true causal variables from false ones. This observation aligns with current studies on environment variable identification for causal inference^21–23^.

We also compared I-GCM with PC-simple in high-dimensional settings. The largest gain was in PPV (Fig. 2b). Because Tables 1–2 and Fig. 2 each reflect a single draw per scenario, we repeated all scenarios over 10 independent replicates, redrawing the DAG, data and environment node each time (Fig. 3). I-GCM achieved a higher PPV than PC-simple in all 10 replicates at every size. Mean PPV over replicates was 0.887 for I-GCM versus 0.659 for PC-simple, a mean gain of 0.21 at 1,000 variables rising to 0.29 at 3,000 (Fig. 3b). Sensitivity was comparable or slightly lower for I-GCM, and F1 and F0.5 were higher for I-GCM, especially for high-dimensional setting(Fig. 3a).

**Fig. 3.**
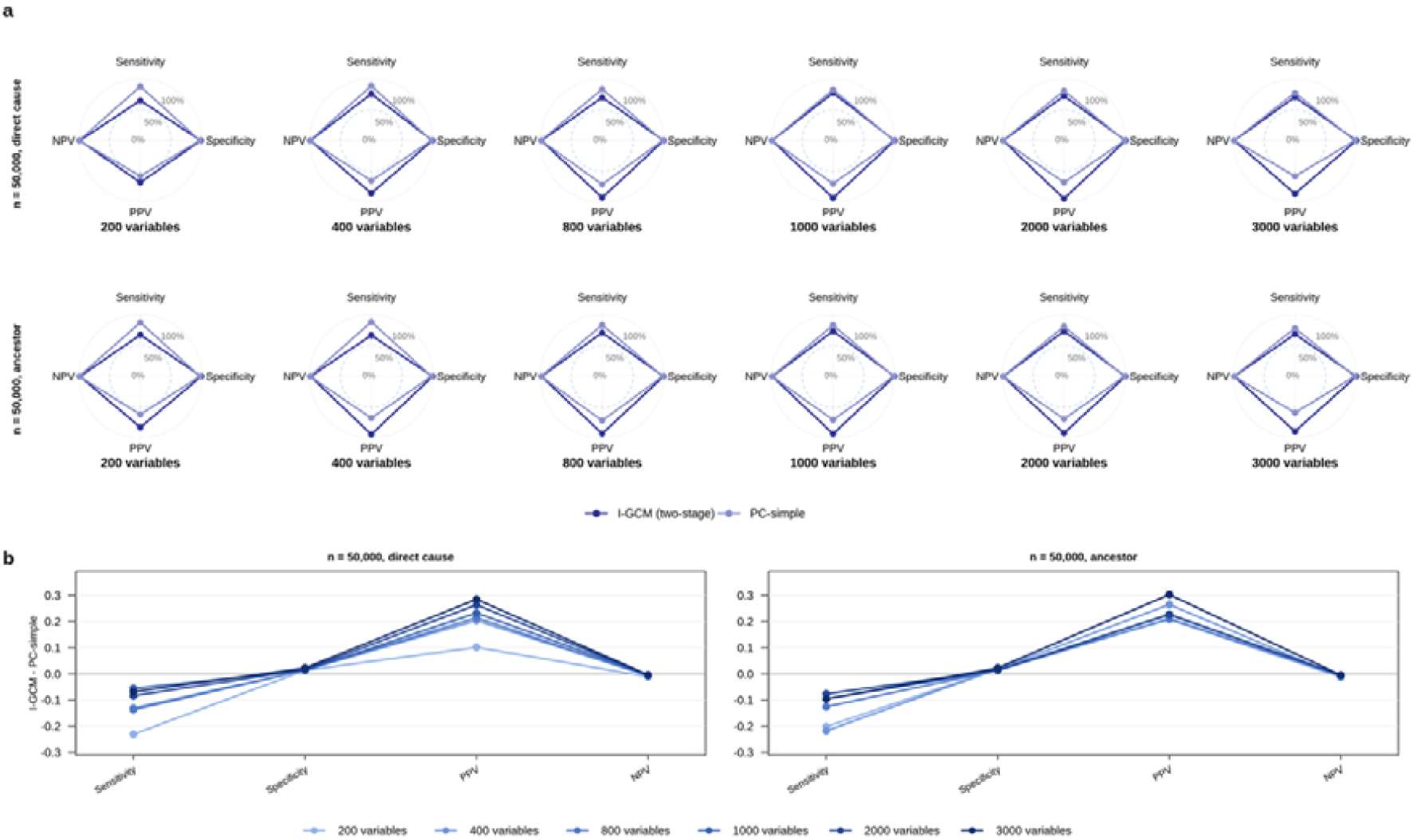
Performance of I-GCM and PC-simple over 10 independent replicates per high-dimensional scenario (n = 50,000). a. Mean PPV, sensitivity, NPV and specificity over replicates for each method. b. Mean difference (I-GCM − PC-simple) in each metric over replicates, by number of input variables.

We evaluated the performance of the PC-simple between a stringent (i.e., α = 0.01) and the default alpha cutoff (i.e., α = 0.05). As anticipated, a decrease in alpha modestly improved the PPV, albeit at the expense of reduced power (refer to Fig. 4.a, Table 3). I-GCM retained its PPV advantage under the stricter pre-screening cutoff (α = 0.01): 0.17 at 2,000 variables and 0.18 at 3,000 (Table 3, Fig. 4b). I-GCM also delivered satisfactory performance in the presence of non-linear relations (Table S2).

**Table 3.** Comparison of the performance between our proposed I-GCM and the PC-simple algorithm under different alpha cutoffs

| **Alpha for PC-simple** | **Environment variable type** | **No. of input variables** | **No. of overall true causal** | **PC-simple** |  |  |  |  |  | **I-GCM** |  |  |  |  |  |
|---|---|---|---|---|---|---|---|---|---|---|---|---|---|---|---|
|  |  |  |  | PPV | Sensitivity | NPV | Specificity | F1-score | F0.5-score | PPV | Sensitivity | NPV | Specificity | F1-score | F0.5-score |
| 0.01 | Direct cause | 2000 | 114 | 0.746 | 0.859 | 0.993 | 0.985 | 0.797 | 0.831 | 0.913 | 0.737 | 0.984 | 0.995 | 0.815 | 0.891 |
| 0.05 | Direct cause | 2000 | 114 | 0.719 | 0.763 | 0.988 | 0.98 | 0.74 | 0.727 | 0.911 | 0.719 | 0.983 | 0.999 | 0.804 | 0.865 |
| 0.01 | Direct cause | 3000 | 163 | 0.81 | 0.705 | 0.983 | 0.99 | 0.754 | 0.861 | 0.99 | 0.607 | 0.978 | 0.999 | 0.746 | 0.961 |
| 0.05 | Direct cause | 3000 | 163 | 0.567 | 0.748 | 0.985 | 0.967 | 0.645 | 0.596 | 0.973 | 0.663 | 0.981 | 0.999 | 0.789 | 0.89 |

**Fig. 4.**
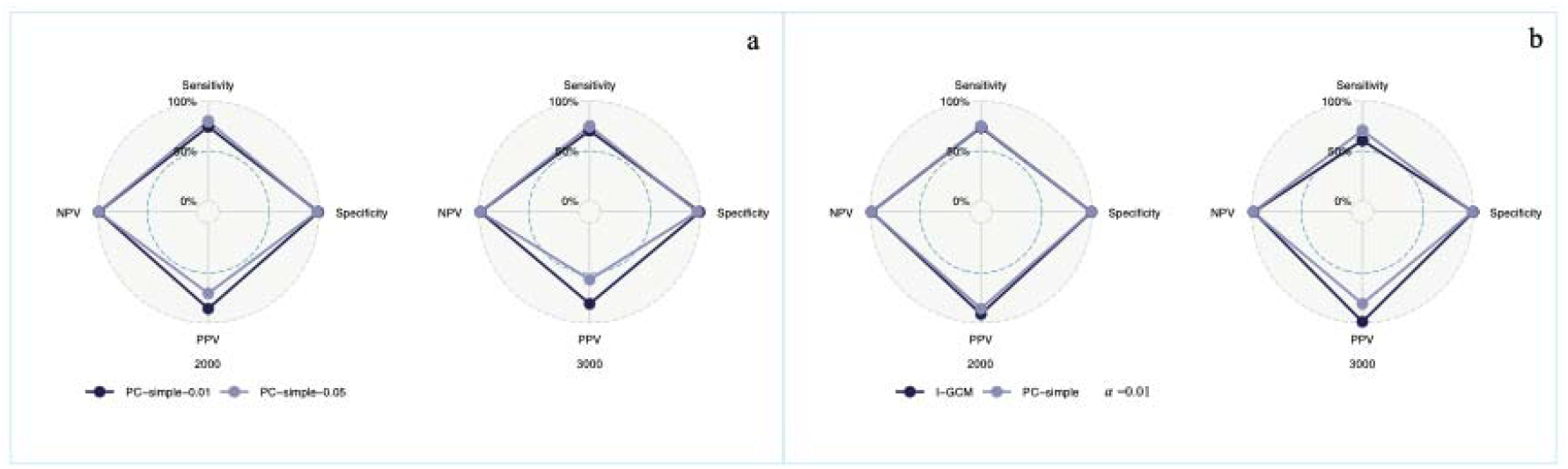
Comparison of causal variable discovery performance across methods and parameter settings. a. Performance of the PC-simple algorithm under different significance cutoffs. b. Comparison between the proposed I-GCM method and the PC-simple algorithm using a stringent cutoff (α = 0.01)

Because sex, the environment variable in our UK Biobank analyses, is likely itself a direct cause of the studied traits, we checked the calibration of the ΔT test in simulations with a direct-cause environment (60 replicates, 3,520 null steps) and with an ancestor environment (40 replicates, 3,154 null steps) in a simulation, applying the selection rule exactly as used (Table S7). Under equation 10, the rejection rate on null steps was close to the nominal level in all tiers but one (α_*ΔT*_ = 0.005, T = 0.1: 0.016 and 0.014), and about 1.3 times nominal overall in both settings (0.026 versus 0.021 expected; 0.018 versus 0.014). Because the same mild excess appeared with an ancestor environment, where *μ* ≈ 0, it reflects slight over-dispersion of the standardized statistic rather than non-centrality. Specifically, each conditioning set differs from the previous one by a single variable, so successive statistics, are positively correlated. Under the null this does not bias individual statistics (their mean remains at zero, i.e. no non-centrality), but it induces slight over-dispersion of the standardized statistics across the sequence, so multiplicity adjustments that assume independence are mildly anticonservative. Because only one variable changes between steps, the effect is small. We observed that employing the non-central variance (Supplementary Text) left the ancestor-environment results unchanged and altered the direct-cause results only slightly (87 versus 93 rejections overall). We therefore report all results under equation 10.

Notably, our proposed method achieved robust performance with the existence of unobserved confounding. It consistently demonstrated superior performance compared to PC-simple(Fig.S1). Most important, the performance, especially PPV, remained robust as the feature dimension increased. As expected, our proposed I-GCM achieved better performance than the association-based method with much higher PPV across simulated scenarios(Fig.S2a). Again, similar pattern was observed with the existence of unobserved confounding(Fig.S2b). We tried different π_*thr*_ threshold for stability selection, our proposed method consistently demonstrated superior performance with much higher PPV across different simulated scenarios.

We also performed a simulation pilot study with stability selection (*p*=1000 candidate variables, *n* = 50000, 50 subsampling iterations at a subsampling fraction of 0.8). Under a π_thr_ threshold of 0.9, on average 48 variables were selected per subsample, giving *E*(*V*) ≈ 3 from Eq. 13 with *p* taken as the full pre-screening count of 1000; as noted in Methods, this bound is heuristic once pre-screening has been applied. Alternatively, if the stability selection procedure is performed on the *entire* algorithm, including the pre-selection step, then the bound is still valid, although this is computationally more demanding. Our model achieved superior performance (PPV=0.963, F1 score=0.932, F0.5 score=0.949) compared with the PC-simple method (PPV=0.706, F1 score=0.807, F0.5 score=0.843). Although this testing scenario involved a moderate feature size, we expect stability selection to demonstrate similar performance in higher-dimensional settings.

### Results for real data application

We next applied I-GCM to large-scale biobank data to demonstrate its ability to yield biologically validated causal insights with clinical implications. Stability selection was not applied to the UK Biobank analyses, which used a single pass of the procedure in Algorithm 1; no formal control of the expected number of false positives is therefore claimed for the biobank results. Table 4 summarizes the number of identified causal variables using “predicted” tissue-specific gene expression levels and clinical variables. The included clinical variables were the same for both COVID-19 infection and severe COVID-19. However, different clinical variables were included for HDL and triglycerides. Only clinical variables relevant to the outcome were included for the causal variables set identification. As illustrated in Table 5, the number of direct causal variables identified for the same target varied when gene expression profiles from different tissues were included. Despite this variation, the identified causal clinical variables remained relatively stable across tissues.

**Table 4.** Identified direct causal variables for different traits.

| **Phenotype** | **No. of total causal variables** | **No. of clinical variables** | **No. of genes** | **Tissue** |
|---|---|---|---|---|
| COVID-19 infection | 36 | 16 | 20 | Whole blood |
| Severe COVID-19 | 90 | 31 | 59 | Whole blood |
| COVID-19 infection | 70 | 17 | 53 | Lung |
| Severe COVID-19 | 156 | 33 | 123 | Lung |
| HDL | 19 | 8 | 11 | Whole blood |
| Triglycerides (TG) | 58 | 16 | 42 | Whole blood |

**Table 5.** Examples of top-ranked (by *Z*_min_) identified direct causal genes for the studied clinical traits.

| **Phenotype** | **Direct causal genes** | **Tissue** |
|---|---|---|
| COVID-19 infection | *CCR5, HLA-DPB2, JAK1, SPR14, RNF138* | Whole blood |
| Severe COVID-19 | *DNLZ, JAK1, PUS10, GPM6A, ORAI1* | Whole blood |
| COVID-19 infection | *FYCO1, KCNN4, VPS16, TULP2, RPUSD4* | Lung |
| Severe COVID-19 | *POM121, PRSA, CCR5, PDE8B, FYCO1* | Lung |
| HDL | *LPL, CCDC92, ACP2, FAM76B, GPR180* | Whole blood |
| Triglycerides (TG) | *BACE1, NRBP1, LPL, TRIM74, SLC56A* | Whole blood |

#### Causal variable identification results for COVID-19 infection

For the blood-specific analysis, 36 (direct) causal variables were identified, while 70 were identified for the lung-specific analysis. Even though the overall causal variables varied between tissues, the direct causal clinical variables were relatively stable across tissues for COVID-19 infection. The identified clinical variables include age, doses of vaccine, Townsend index, BMI, LDL, ethnic group, coronary artery disease (CAD), dementia, atrial fibrillation (AF), and so on (for full details, please refer to Table S3a). Most of these clinical variables have previously been reported to be associated with COVID-19 infection. For example, age has been identified as the most important causal variable for COVID-19 infection both for the analyses with whole blood and the lung. Existing studies have demonstrated that older people, especially those over 70, are more susceptible to severe COVID-19 infection than young adults^24,25^. However, while previous research has focused primarily on correlational associations, this study applies a causal inference framework to identify *direct clinical and genetic causal drivers* for COVID-19.

The identified direct causal genes for COVID-19 infection differed across tissues. Specifically, 20 direct causal genes were identified in whole blood and 53 in the lung. Interestingly, we found that the ranking of identified causal genes varied between tissues. This aligned with our expectation, as COVID-19 is a lung-related disease, and many disease-related genes are tissue-specific. We compared the performance of our method with the PC-simple algorithm in identifying potential targets using the Fisher’s exact test(Table S4). I-GCM performed comparably with PC-simple, although it tended to identify fewer variables. Many of the identified genes have been reported to be COVID-19-associated by existing studies, with some identified as potential drug targets for COVID-19 (Table 5, full results please refer to Table S3a), e.g., *CCR5, FYCO1, KCNN4*, etc. Patterson et al.^26^ reported that inhibiting *CCR5* in COVID-19 patients could lead to decreased inflammatory cytokines and COVID-19 RNA in the plasma. *FYCO1* has been identified as a COVID-19-associated gene from several GWAS^27,28^. Specifically, abnormal expression of this gene may lead to respiratory failure in COVID-19 patients^29^.

As expected, many of the enriched pathways have been reported to be relevant to COVID-19 or related pathophysiology (Table S5a). Here we highlighted a few interesting pathways. As one of the top enriched pathways for COVID-19 infection, the N-acetylglucosamine (NAG) degradation pathway has been reported to be involved in chronic anti-inflammatory reactions, and a clinical study reported that NAG may lead to reduced hospital stay and ICU admission for COVID-19 patients^30^. Acute viral myocarditis was another top-enriched pathway. Notably, acute myocarditis has been reported as a complication of COVID-19 infection ^31^. The mitochondrial transcription initiation pathway, significantly enriched for COVID-19 infection, has been reported to be associated with the mediation effects of COVID-19 on innate immunity^32^.

Drug enrichment analysis identified no compound passing multiple-testing correction for COVID-19 infection (Table S6a). Among nominally enriched compounds, cyclosporin A ranked second (p = 1.2 × 10C^3^), consistent with a role for calcineurin-dependent immune signaling. However, the results require experimental validations, and we draw no therapeutic inference from these exploratory results.

#### Causal variable identification results for severe COVID-19

For severe COVID-19, we identified 90 and 156 (direct) causal variables for the whole blood-specific and lung-specific analyses, respectively. Again, the identified clinical causal variables remained stable across tissues (i.e., whole blood and lung). The identified direct clinical variables include doses of vaccine, stroke, Townsend index, non-COVID-19 pneumonia, type 2 diabetes mellitus (T2DM), renal failure, chronic obstructive pulmonary disease (COPD), heart failure, venous thrombus embolism (VTE), waist circumference, HDL etc. (full results please refer to Table S3b). All identified clinical variables have been reported as risk factors for COVID-19 severity in existing studies, however, here we further demonstrate they are likely *direct causal* factors.For example, two doses of COVID-19 vaccine was identified as the most important (ranked by *Z*_*min*_) direct causal variable from our study. This finding aligns with existing studies which showed that COVID-19 vaccination is highly effective in preventing hospital admission/death from COVID-19. A meta-analysis on 7,267,055 COVID-19 patients revealed that stroke could lead to increased COVID-19 mortality with a pooled effect estimate of 1.3^33^. T2DM was also one of the earliest identified risk factors for COVID-19 severity^34^. COVID-19 patients with T2DM have been reported to demonstrate poor therapeutic responses^35^. Other identified causal variables like stroke, non-COVID-19 pneumonia, Townsend index, renal failure, and COPD were also well-acknowledged risk factors for COVID-19 severity.

We respectively identified 59 and 123 direct causal genes for the whole blood and the lung (Table S3b). Similar to COVID-19 infection, the rankings of the identified direct causal genes were highly tissue-specific. To evaluate their potential as therapeutic targets, we compared our prioritized genes against those selected by the PC-simple algorithm using target annotations from the OpenTargets database as the standard.. We conducted Fisher’s exact test(Table S4) to examine whether there exist significant detection power differences between our method and the PC-simple algorithm. Comparable power was observed even though fewer causal genes were identified by our method. In other words, the sensitivity for our method is not significantly different from that for PC-simple algorithm based on this empirical comparison. Encouragingly, many of the identified genes have been reported to be associated with severe COVID-19 in existing studies, e.g., *DNLZ, JAK1, CCR5, PDE8B, POM121*, etc. (Table 5; please refer to Table S3b for more details). For example, Andreini et al.^36^ reported that *DNLZ* could affect the binding of zinc(II), which may lead to inefficient immune responses to COVID-19 infection. Chen et al.^37^ revealed that inhibition of JAK1 could ameliorate cytokine release syndrome. It is worth noting that baricitinib, a treatment that targets JAK1, is used to treat COVID-19 in clinical practice^38^. By leveraging ICP across tissue-specific biobank data, our approach moves beyond traditional pairwise associations and instrumental variable assumptions to establish robust, *direct causal evidence* supporting *JAK1* as a therapeutic target in severe COVID-19. These findings also demonstrate that identifying direct causal genes may significantly enhance drug discovery or repositioning efforts.

We also performed pathway enrichment analysis on the identified causal gene set (Table S5b). Many of the enriched pathways were significantly associated with the pathophysiology of COVID-19 or related complications. For example, ‘other interleukin signaling’ was one of the top-enriched pathways for severe COVID-19. Interleukin signaling pathways were intensively involved in anti-inflammatory activity ^39^, and thus may help the early identification of severe COVID-19 patients. Membrane trafficking was also identified as a top-enriched pathway. As suggested by Banerjee et al.^40^, interrupted membrane protein trafficking could lead to the disruption of signal recognition particles, which in turn could promote the propagation of COVID-19. Vesicle-mediated transport was another significantly enriched pathway. In a previous study by Hassanpour et al.^41^, exosomes (an extracellular vesicle) may play a role in COVID-19 virus infection.

Drug enrichment analysis for severe COVID-19 likewise identified no compound passing multiple-testing correction (Table S6b). Cyclosporin A was the only agent nominally enriched in both the infection and severity analyses.

#### Causal variable identification results for HDL

In total, we identified 19 direct causal variables for HDL, comprising 8 clinical variables and 11 genes (Table S3c). Four of the selected clinical variables, apolipoprotein A, triglycerides, total cholesterol and LDL, are measured from the same blood draw as the outcome and are compositionally related to it: apolipoprotein A is the principal structural protein of the HDL particle, and total cholesterol mathematically contains HDL-C. Their selection reflects this compositional coupling rather than evidence of a causal effect, and we do not interpret them causally.

Among the remaining clinical variables, waist–hip ratio, BMI and hip circumference are established anthropometric correlates of HDL level, consistent with the role of adiposity in HDL metabolism. Haemoglobin (Hb) has been linked to HDL particle size by Hämäläinen et al. ^42^, who reported that a higher plasma Hb concentration is associated with larger HDL particles, which in turn may be associated with increased risk of metabolic and cardiovascular conditions. While clinical risk factors for lipid abnormalities are widely recognized, existing literature has predominantly focused on observational associations. A central contribution of this study is to isolate direct causal drivers from correlational markers.

Furthermore, many identified causal genes are supported by previous studies (Table 5, full results please refer to Table S3c). For example, Xiao et al.^43^ reported that *CCDC92* has an effect on the size and concentration of HDL particles in plasma. As suggested by Tsutsumi et al.^44^, a decrease in LPL activity was associated with unfavorable HDL levels in the plasma.

Pathway enrichment analysis was also performed for HDL (Table S5c). Some top pathways include ‘composition of lipid particles’, ‘metabolic pathway of LDL, HDL and TG, including diseases’, and ‘role of ppar-gamma coactivators in obesity and thermogenesis’. Kersten^45^ reported that the activation of PPAR receptors could lead to an increased HDL level in plasma. For more details about the enriched pathways, please refer to Table S5c.

#### Causal variable identification results for TG

For TG, we identified a total of 58 direct causal variables, with 16 clinical variables and 42 genes (Table S3d). As in the HDL analysis, several of the selected clinical variables, HDL, apolipoprotein A, apolipoprotein B, lipoprotein(a) and total cholesterol, derive from the same lipid panel as the outcome and are interpreted as compositional or metabolic correlates rather than direct causes.

The remaining clinical variables are consistent with established metabolic determinants of triglyceride level. Glucose and glycated haemoglobin (HbA1c) reflect glycemic control, and Srinivasan et al.^46^ reported that poor glucose metabolism is associated with elevated triglycerides; type 2 diabetes and hypertension are components of the metabolic syndrome, of which hypertriglyceridemia is a defining feature; and smoking status, ethnicity and medication use are recognized correlates of circulating triglyceride level.

We note that triglycerides were selected in the HDL analysis and HDL in the triglyceride analysis. Under the acyclicity assumption underlying our framework, two variables cannot each be a direct cause of the other. This mutual selection therefore indicates that the two traits are strongly coupled compositional and metabolic correlates measured contemporaneously, rather than that either causes the other, and illustrates a general limitation of applying invariance-based causal discovery to variables drawn from a single measurement panel.

Notably, many identified genes are also supported by the literature (Table 5, full results please refer to Table S3d). For example, Meakin et al.^47^ suggested that the loss of BACE1 could lead to unfavorable lipid levels. *NRBP1* has been identified as a susceptibility gene for TG based on an independent GWAS by Read et al.^48^.

Table S5d demonstrates the pathway enrichment analysis result for TG. Here we highlight a few top pathways. Integrin signaling pathway was one of the top-enriched pathways based on the identified causal gene set. In a related study, Xiao et al.^49^ demonstrated that integrin β3 deficiency was associated with elevated triglyceride levels. Cholesterol metabolism was another significantly enriched pathway for TG. According to Feingold^50^, the removal of triglycerides in very low-density lipoprotein was associated with cholesterol levels. NRF2 pathway was also found to be significantly enriched for TG. In a study by Tanaka et al.^51^, the NRF2 pathway was shown to inhibit the accumulation of triglycerides in the blood.

## Discussion

This study introduced a novel methodological framework, I-GCM, to identify direct causal variables for the target of interest. The method combines the GCM with the invariance property of causal relationships for effective causal variables discovery. Our work establishes a practical path for causal discovery in biobank-scale data, bridging the gap between statistical causality and translational medicine, while maintaining computational feasibility. Simulation results validated the efficacy of the proposed method in detecting actual causal variables. Across replicated high-dimensional simulations, I-GCM raised PPV by 0.21–0.29 relative to PC-simple while maintaining comparable sensitivity (Fig. 3).

Also, we applied our proposed approach to 2 binary and 2 continuous traits extracted from the UK Biobank (i.e., COVID-19 infection, severe COVID-19, HDL-C, and TG) to uncover direct causal clinical and genetic variable-sets in the whole blood and the lung. Encouragingly, most of the identified clinical causal variables are supported by the literature. Additionally, we found that gene sets identified by our method were significantly enriched in pathways involved in the pathophysiology of the studied traits. Given the satisfactory performance of I-GCM in identifying causal variables, this framework may be particularly useful for prioritizing genes for experimental follow-up studies. Furthermore, the identified causal genes may serve as potential targets for novel treatments and drugs.

The proposed framework I-GCM has several strengths. Its main advantage is precision: relative to PC-simple alone, I-GCM reduced false positives substantially at comparable sensitivity, and the gain grew with the number of candidate variables. Importantly, we showed that integrating structural causal discovery methods (e.g. PC algorithm-based methods) with the more recently proposed invariance-based methods (ICP) may improve both causal discovery performance and computational speed. This advancement opens up possibilities for applying ICP methods to high-dimensional data.

Another strength of I-GCM is its ability to capture both linear and non-linear relationships, eliminating the need for prior knowledge about the underlying generative model. We employed the GCM as a measure of dependence, which allows non-linear relationship between the covariates and the outcome or environment. Many existing or common causal inference approaches, such as PC/PC-simple and the original ICP approach, assume linear relationships between the variables. Although we utilized gradient boosted trees (XGBoost) in our study, any machine learning methods, including deep neural networks, could be employed to calculate GCM to detect the relations between variables and the target. Our method can also potentially combine datasets collected from different experimental settings (e.g., observational/interventional datasets) and leverage them for causal discovery. This makes our method a natural fit for datasets from different sources, data obtained under different interventions (e.g. knockout or perturbation of different genes, different medication treatment, different experimental conditions etc.), or data obtained under both interventional and observational settings. However, it may be challenging to use the PC-simple algorithm per se in such situations, as it is not straightforward how the different kinds of data can be combined together.

Furthermore, performing variable selection before causal variable discovery substantially reduces computation complexity. Since we ranked the preselected variables and used a backward feature selection method to enumerate the candidate causal variable sets, we dramatically reduce the number of candidate sets (*q vs* 2^q^), leading to massively improved computational speed (Table S8). To the best of our knowledge, this study is among the first to exploit invariant causal prediction in genetic epidemiology studies, and the first to apply ICP principles in biobank-scale datasets with high-dimensional genomic data. Given the increasing availability of large-scale datasets, our method shows great potential for identifying reliable (direct) causal variables for diseases. The identified causal variables could provide insights into underlying disease mechanisms and inform more effective treatment and prevention strategies.

There are several limitations to this study. The PC-simple pre-screen targets linear dependence and may miss purely non-linear associations, although our simulations indicate that I-GCM performs reasonably when such relations are present. The GCM test is valid asymptotically and assumes sufficiently fast convergence of the conditional-mean estimators; we used gradient-boosted trees without sample splitting, so residual estimation error is not formally controlled. The variance in equation 10 is exact only when the environment variable is not a direct cause of the outcome; when it is, as for sex in our UK Biobank analyses, the statistics are non-central and we rely on simulation evidence of calibration (Table S7), obtained with a continuous environment dichotomized at its median rather than with sex. Nevertheless, we also presented the non-central version in the supplementary text. The standardized ΔT statistic was mildly over-dispersed under the null (SD ≈ 1.1), and because the selection rule keys on a single change point, small changes in the variance may still alter the selected set in a minority of cases (7 of 131 replicates when the non-central form was substituted(Table S7)).

The change-point statistic and the backward elimination over nested candidate sets are considered heuristics; the search is greedy and the prioritized feature sets carry no formal guarantee of global optimality. The identified sets are also conditional on the environment variable: in exploratory analyses with alternative environments (e.g., ancestry PC1), the top-ranked clinical variables were selected consistently, while the number of selected genes varied with the environment. Gene-level findings should therefore be considered as specific to the sex environment. We model only single-directional relations, although reverse causality is unlikely for genotype-predicted expression and covariates were restricted to measurements preceding the outcome. Our lipid analyses illustrate a further limitation: fractions measured on the same assay panel are related by construction (total cholesterol contains HDL-C, and apolipoprotein A is a structural component of the HDL particle) so no invariance-based criterion can distinguish a causal from a definitional relation. Consistent with this, triglycerides and HDL were each selected in the other’s analysis, which cannot hold under acyclicity; we therefore restrict causal interpretation of the lipid traits to non-panel variables and the identified genes.

As for the COVID-19 analyses, they were restricted to tested participants. Also, the controls of the severe COVID-19 analysis combined uninfected and non-hospitalized infected individuals, so these results reflect both susceptibility to infection and progression given infection. Ancestry principal components were not included among the conditioning variables, so some gene associations may reflect residual population structure. For our application scenarios, the cohort is close to ancestrally homogeneous. Finally, UK Biobank participants are healthier and less socioeconomically diverse than the general population^52^, which limits generalizability.

In conclusion, we have proposed a novel and scalable framework for causal variable discovery with high precision. Our proposed framework is flexible and could be extended to other types of omics data, such as methylation, scRNA-seq and scATAC-seq. Beyond methodological innovation, our study reveals biologically meaningful direct causal determinants of major traits. I-GCM identified biologically important and potentially druggable causal targets for COVID-19 and lipid abnormalities, such as *JAK1, CCR5 and LPL*, offering new hypotheses for therapeutic modulation. As biomedical data continue to grow in scale and complexity, frameworks such as I-GCM will be essential for bridging statistical causality and clinical translation. Future extensions to multi-omics and longitudinal data could accelerate the identification of disease mechanisms and drug targets, paving the way for precision therapeutics.

## Supporting information

Ver3.9h_SupplementaryFile

## Data Availability

UK Biobank data are not publicly available but can be accessed by researchers through a formal application process. Eligibility criteria and application details are available on the UK Biobank website: https://www.ukbiobank.ac.uk/use-our-data/. The source codes and R package to reproduce our experiments are available via Github at https://github.com/LiangyingYin/I-GCM.

## Data availability

UK Biobank data are not publicly available but can be accessed by researchers through a formal application process. Eligibility criteria and application details are available on the UK Biobank website: https://www.ukbiobank.ac.uk/use-our-data/.

## Code availability

The source codes and R package to reproduce our experiments for this work are available via Github at https://github.com/LiangyingYin/I-GCM.

## Acknowledgements

This work was supported partially by a Young Collaborative research grant (C4003-23Y), a National Natural Science Foundation China grant (81971706), the Lo Kwee Seong Biomedical Research Fund from The Chinese University of Hong Kong and the KIZ-CUHK Joint Laboratory of Bioresources and Molecular Research of Common Diseases, Kunming Institute of Zoology and The Chinese University of Hong Kong, China. An earlier version of this manuscript was posted on MedRxiv:

