## Supplementary material for "A scalable invariant causal discovery framework reveals direct genetic and clinical determinants of complex human traits": Ver3.9h_SupplementaryFile

**Supplementary Text**

**Detailed derivation for equation 10:**

Var ($\Delta T_{S_{j-1},S_{j}}^{n})=Var(\left| T_{S_{j-1}}^{n} \right|-\left| T_{S_{j}}^{n} \right|)$

$=Var\left( \left| T_{S_{j-1}}^{n} \right| \right)+Var\left( \left| T_{S_{j}}^{n} \right| \right)-2Cov\left( \left| T_{S_{j-1}}^{n} \right|,\left| T_{S_{j}}^{n} \right| \right)$

$$=Var\left( \left| \left| T_{S_{j-1}}^{n} \right| \right| \right)+Var\left( \left| T_{S_{j}}^{n} \right| \right)-2\left[ E\left( \left| T_{S_{j-1}}^{n}*T_{S_{j}}^{n} \right| \right)-E\left( \left| T_{S_{j-1}}^{n} \right| \right)*E\left( \left| T_{S_{j}}^{n} \right| \right) \right]=1-\frac{2}{\pi}+ 1-\frac{2}{\pi}-2*\left[ \frac{2}{\pi}\left( \rho*arcsin\rho+\sqrt{1-\rho^{2}} \right)-\frac{2}{\pi} \right]$$

$=2-\frac{4}{\pi}(\rho*arcsin\rho+ \sqrt{1-\rho^{2}})$  (S1)

where $\rho$ indicates the correlation between $T_{S_{j-1}}^{n}$ and $T_{S_{j}}^{n}$. Since both $T_{S_{j-1}}^{n}$ and $T_{S_{j}}^{n}$ follow standard normal distributions under the null, the correlation between $T_{S_{j-1}}^{n}$ and $T_{S_{j}}^{n}$ also equals the covariance between them, i.e., $\rho=Cor\left( T_{S_{j-1}}^{n},T_{S_{j}}^{n} \right)=Cov(R_{S_{j-1}}^{n},R_{S_{j}}^{n})/(se\left( R_{S_{j-1}}^{n} \right)*se\left( R_{S_{j}}^{n} \right)*N))$. Note that under the null hypothesis, we assume that *E* is independent of *Y* given the set of covariates (which include the set of direct causes), and that removing the *j^t^*^h^ variable does not affect this independence as the *j^t^*^h^ variable is not a direct cause itself. Following the above argument, both $\left| T_{S_{j-1}}^{n} \right|$ and $\left| T_{S_{j}}^{n} \right|$ follow folded normal distribution. Following the definition for folded normal distribution, the variance and mean can be respectively represented as follows^1^ :

$Var\left( {|T}_{S_{j-1}}^{n}| \right)= \mu^{2}+ \sigma^{2}-{[\sigma\sqrt{\frac{2}{\pi}}e^{(-\mu^{2}/2\sigma^{2})}+ \mu(1-2\Phi(-\frac{\mu}{\sigma}))]}^{2}=1-\frac{2}{\pi}= \sigma_{T_{S_{j-1}}^{n}}^{2}$  (S2)

$E\left( {|T}_{S_{j-1}}^{n}| \right)=\sigma\sqrt{\frac{2}{\pi}}e^{(-\mu^{2}/2\sigma^{2})}+ \mu(1-2\Phi(-\frac{\mu}{\sigma}))= \sqrt{\frac{2}{\pi}}=\mu_{T_{S_{j-1}}^{n}}$  (S3)

where $\mu$ and $\sigma$ respectively indicates the mean and standard deviation of $T_{S_{j-1}}^{n}$, i.e., 0 and 1. This also applies for $Var\left( \left| T_{S_{j}}^{n} \right| \right)$. Following Shojaie et al. ^1^, we have:

$E\left( \left| T_{S_{j-1}}^{n}*T_{S_{j}}^{n} \right| \right)=\frac{2\sigma_{T_{S_{j-1}}^{n}}* \sigma_{T_{S_{j}}^{n}}}{\pi}(\rho*arcsin\rho+ \sqrt{1-\rho^{2}})$  (S4)

**Extension to the case where the environment variable is a direct cause of the target (non-central case)**

Equation 10 of the main text assumes that, under the null hypothesis, *E* ⊥ *Y* | *S* holds exactly, so that both GCM statistics have mean μ = 0 and are standard normal. When *E* is itself a direct cause of *Y*, as for sex in the UK Biobank analyses, the conditional dependence between *E* and *Y* given the remaining direct causes is invariant but non-zero. Under the null hypothesis that the *j*-th variable is not a direct cause, the two statistics then share a common non-zero mean *μ* and unit variance, and ($T_{S_{j-1}}^{n}$, $T_{S_{j}}^{n}$) is asymptotically bivariate normal with means (*μ, μ*), unit variances and correlation *ρ* given by equation 11. Evaluating the folded-normal moments (S2) and (S3) at this *μ* with σ = 1 gives $E\left| T \right|=\sqrt{2/\pi} e^{-\mu^{2}/2}+\mu\left[ 1-2\Phi(-\mu) \right]$ and $\text{Var}\left| T \right|=\mu^{2}+1-\left( E\left| T \right| \right)^{2}$, and the cross-moment (S4) becomes

$E\left| T_{S_{j-1}}^{n} T_{S_{j}}^{n} \right|=\iint\left| t_{1}t_{2} \right| \varphi_{2}\left( t_{1},t_{2};\mu,\mu,\rho\right) dt_{1} dt_{2}$  (S5)

where φ₂ denotes the bivariate normal density with means (*μ, μ*), unit variances and correlation ρ. A closed-form expression for (S5) is given by Haas et al.²; in our implementation we evaluate it by exact one-dimensional conditional integration. The implementation reproduces equation 10 at μ = 0 (difference < 10⁻¹⁰), tends to 2(1 − ρ) for |μ| ≥ 5 (difference ≤ 2×10⁻⁶) and agrees with Monte Carlo evaluation (10⁷ draws) to within 1.4×10⁻⁴. Substituting these moments into the second line of (S1) gives

$\text{Var}\left( \Delta T^{n} \right)=2 \text{Var}\left| T \right|-2\left[ E\left| T_{S_{j-1}}^{n} T_{S_{j}}^{n} \right|-\left( E\left| T \right| \right)^{2} \right]$  (S6)

which reduces to equation 10 when *μ* = 0 and to 2(1 − *ρ*) as |μ| → ∞, where |T| ≈ T. In practice *μ* is unknown and is replaced by the plug-in estimate $\hat{\mu}=T_{S_{j}}^{n}$, the statistic computed on the larger of the two nested sets, which shares the same mean under the null. Because consecutive nested sets differ by a single variable, *ρ* is close to one (median 0.997 across simulation runs), and the non-central variance exceeded equation 10 by a median of 3–5% (at most only ~13%). The empirical calibration of the ΔT test under both variance forms is reported in Table S7; results in the main text use equation 10, as the results are very similar regardless of using equation 10 or the non-central counterpart.

**Supplementary Tables**

Table S1 Summary of studied phenotypes in UK-Biobank

| Phenotype | Sample size | No. of covariates | Tissue |
| --- | --- | --- | --- |
| COVID-19 infection | 154749 | 6366 | Whole blood |
| Severe COVID-19 | 154749 | 6366 | Whole blood |
| COVID-19 infection | 154749 | 8037 | Lung |
| Severe COVID-19 | 154749 | 8037 | Lung |
| HDL-C | 328002 | 6349 | Whole blood |
| Triglycerides (TG) | 328002 | 6349 | Whole blood |

Table S2 Simulation results for our proposed I-GCM and the PC-simple algorithm with both linear and non-linear causal relations

Table S3 Identified direct causal variables for studied traits

Table S4 Comparison of I-GCM and the PC-simple algorithm in revealing potential gene targets from the OpenTargets database

| Trait | Tissue | Odds Ratio | P-value |
| --- | --- | --- | --- |
| COVID-19 infection | Whole blood | 1.67 | 0.87 |
| Severe COVID-19 | Whole blood | 0.5 | 0.46 |
| COVID-19 infection | Lung | 1.35 | 0.79 |
| Severe COVID-19 | Lung | 1.09 | 0.68 |

Note: a p-value > 0.05 indicates that no significant difference between the two methods in revealing potential targets was detected from the OpenTargets database

Table S5 Pathway enrichment analysis result for studied traits

Table S6 Drug enrichment analysis result for COVID infection and severe COVID

Table S7 Empirical calibration of the ΔT test in simulations with a direct-cause or an ancestor environment variable

| Tier (screened set size q) | α; threshold T | Replicates | Null steps | Expected rejections (α × null steps) | Null steps rejected, eq. 10 | Null steps rejected, non-central | Power, eq. 10 | Power, non-central |
| --- | --- | --- | --- | --- | --- | --- | --- | --- |
| Direct-cause environment variable (60 replicates) | | | | | | | | |
| q ≤ 60 | 0.05; 0 | 40 | 1,209 | 60.5 | 0.060 (73) | 0.058 (70) | 0.75 (9/12) | 0.75 (9/12) |
| 60 < q ≤ 110 | 0.01; 0 | 4 | 305 | 3.1 | 0.007 (2) | 0.007 (2) | 0.67 (4/6) | 0.50 (3/6) |
| 110 < q ≤ 160 | 0.005; 0.1 | 6 | 560 | 2.8 | 0.016 (9) | 0.013 (7) | 0.33 (2/6) | 0.33 (2/6) |
| 160 < q < 210 | 0.005; 0.2 | 8 | 1,135 | 5.7 | 0.008 (9) | 0.007 (8) | 0.47 (8/17) | 0.47 (8/17) |
| q ≥ 210 | 0.001; 0.3 | 2 | 311 | 0.3 | 0.000 (0) | 0.000 (0) | 0.25 (1/4) | 0.25 (1/4) |
| All tiers | as applied | 60 | 3,520 | 72.3 | 0.026 (93) | 0.025 (87) | 0.53 (24/45) | 0.51 (23/45) |
| Ancestor environment variable (40 replicates) | | | | | | | | |
| q ≤ 60 | 0.05; 0 | 19 | 643 | 32.1 | 0.058 (37) | 0.058 (37) | 0.71 (12/17) | 0.71 (12/17) |
| 60 < q ≤ 110 | 0.01; 0 | 4 | 279 | 2.8 | 0.014 (4) | 0.014 (4) | 0.62 (5/8) | 0.62 (5/8) |
| 110 < q ≤ 160 | 0.005; 0.1 | 7 | 701 | 3.5 | 0.014 (10) | 0.014 (10) | 0.29 (4/14) | 0.29 (4/14) |
| 160 < q < 210 | 0.005; 0.2 | 8 | 1,189 | 5.9 | 0.005 (6) | 0.005 (6) | 0.68 (17/25) | 0.68 (17/25) |
| q ≥ 210 | 0.001; 0.3 | 2 | 342 | 0.3 | 0.000 (0) | 0.000 (0) | 0.00 (0/5) | 0.00 (0/5) |
| All tiers | as applied | 40 | 3,154 | 44.7 | 0.018 (57) | 0.018 (57) | 0.55 (38/69) | 0.55 (38/69) |

Notes: simulations with n = 50,000 and 10 replicates per p (p = 200–3,000 with a direct-cause environment; 400–3,000 with an ancestor environment). Each replicate falls into one tier of the adaptive rule according to its screened set size q, and a backward step is rejected when p ≤ α and |ΔT| > T; “All tiers” pools the replicates under the rule as applied. Steps were classified from the data-generating DAG: null if removing the variable changes the population mean of ΔT by < 0.05, non-null if by ≥ 0.2. Rejection rate = proportion of null steps rejected (number in parentheses); power = proportion of non-null steps rejected (rejected/total), imprecise because non-null steps are few. Eq. 10 and non-central denote the two forms of Var(ΔT): under μ = 0, and with the plug-in mean (equation S6); for the ancestor environment both give identical results. Steps within a replicate are not independent, so no confidence intervals are given.

Table S8 Wall-clock runtime of I-GCM and invariance-test counts compared with nonlinear ICP

| p | n | q, mean (range) | PC-simple (s) | I-GCM nested loop (s) | I-GCM total (s) | ICP subsets (2^q^) | Nonlinear ICP (s) |
| --- | --- | --- | --- | --- | --- | --- | --- |
| 200 | 50,000 | 15.3 (10–20) | 2 ± 0 | 33 ± 14 | 42 ± 15 | 4.0 × 10^4^ | 4,209 (q = 10); 42,665 (q = 13)^a^ |
| 400 | 50,000 | 25.1 (17–31) | 2 ± 0 | 61 ± 33 | 89 ± 35 | 3.6 × 10^7^ | ≥ 1.5 × 10^8^ (≈ 4.7 years)^b^ |
| 800 | 50,000 | 47.8 (39–54) | 12 ± 4 | 308 ± 38 | 411 ± 41 | 2.5 × 10^14^ | ≥ 1.0 × 10^15^ (≈ 3 × 10^7^ years)^b^ |
| 1,000 | 50,000 | 54.1 (49–60) | 18 ± 6 | 335 ± 77 | 508 ± 96 | 1.9 × 10^16^ | ≥ 7.9 × 10^16^ (≈ 3 × 10^9^ years)^b^ |
| 2,000 | 50,000 | 115.2 (100–128) | 388 ± 126 | 1,620 ± 259 | 2,610 ± 322 | 4.8 × 10^34^ | ≥ 2.0 × 10^35^ (≈ 6 × 10^27^ years)^b^ |
| 3,000 | 50,000 | 193.3 (161–215) | 5,051 ± 1,697 | 4,685 ± 814 | 11,104 ± 2,298 | 1.5 × 10^58^ | ≥ 6.3 × 10^58^ (≈ 2 × 10^51^ years)^b^ |
| 1,000 | 30,000 | 53.6 (45–64) | 15 ± 5 | 172 ± 64 | 276 ± 72 | 1.4 × 10^16^ | ≥ 5.6 × 10^16^ (≈ 2 × 10^9^ years)^b^ |
| 2,000 | 30,000 | 104.9 (94–124) | 250 ± 102 | 714 ± 162 | 1,289 ± 230 | 3.8 × 10^31^ | ≥ 1.6 × 10^32^ (≈ 5 × 10^24^ years)^b^ |
| 3,000 | 30,000 | 171.4 (142–190) | 2,239 ± 664 | 1,884 ± 380 | 4,878 ± 1,023 | 3.9 × 10^51^ | ≥ 1.6 × 10^52^ (≈ 5 × 10^44^ years)^b^ |

Note: n, sample size; p, number of input variables; q, number of variables retained after PC-simple screening. Runtimes are wall-clock seconds, mean ± SD over 10 replicates (direct-cause environment); PC-simple used 8 and the I-GCM nested loop 14 parallel workers (AMD Ryzen Threadripper 7960X, 251 GB RAM; R 4.3.1), and I-GCM total also includes data generation. I-GCM performs q − 1 invariance tests, whereas ICP must test all 2^q^ subsets (shown for the mean q). ^a^Measured: nonlinear ICP (nonlinearICP 0.1.2.1, invariant residual distribution test, α = 0.05) was run single-threaded on the same screened variables and data, with the environment dichotomised at its median. It accepted no subset and returned the empty set in both runs, as expected when the environment is a direct cause of the outcome; on the same data I-GCM selected 6 variables, all true parents, at q = 10 (PPV 1.00, sensitivity 0.60; 83 s single-threaded) and abstained at q = 13. ^b^Projected as 2^q^ × 4.11 s per subset, the rate measured at q = 10; these are lower bounds because the per-subset cost grows with q.

**Supplementary Figures**

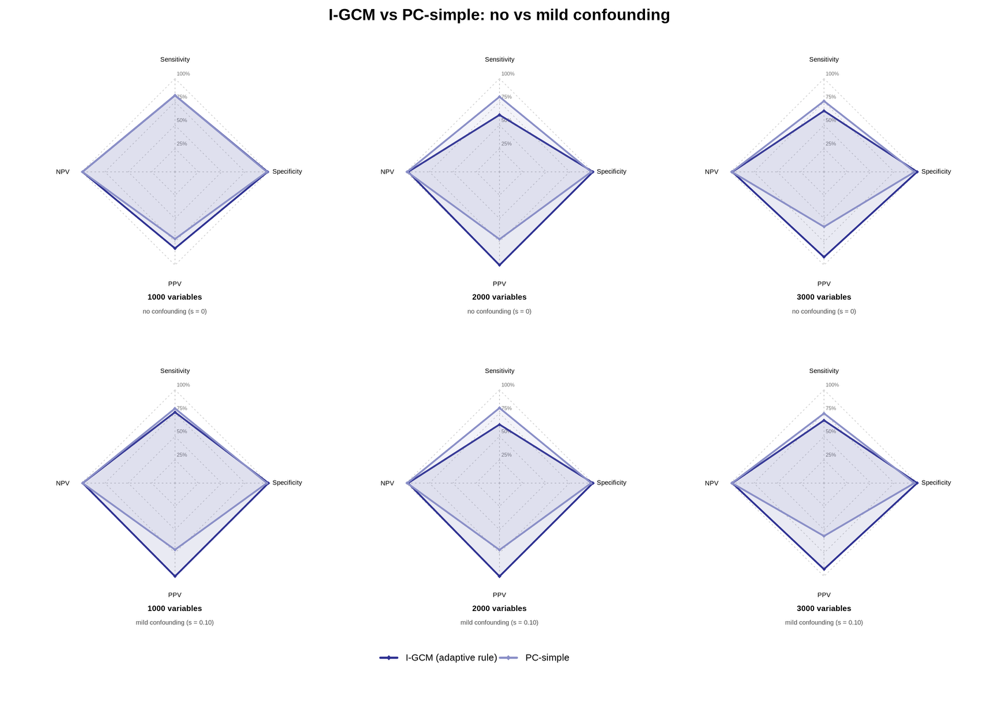

Fig.S1 Performance comparison between without and with unobserved confounding scenarios

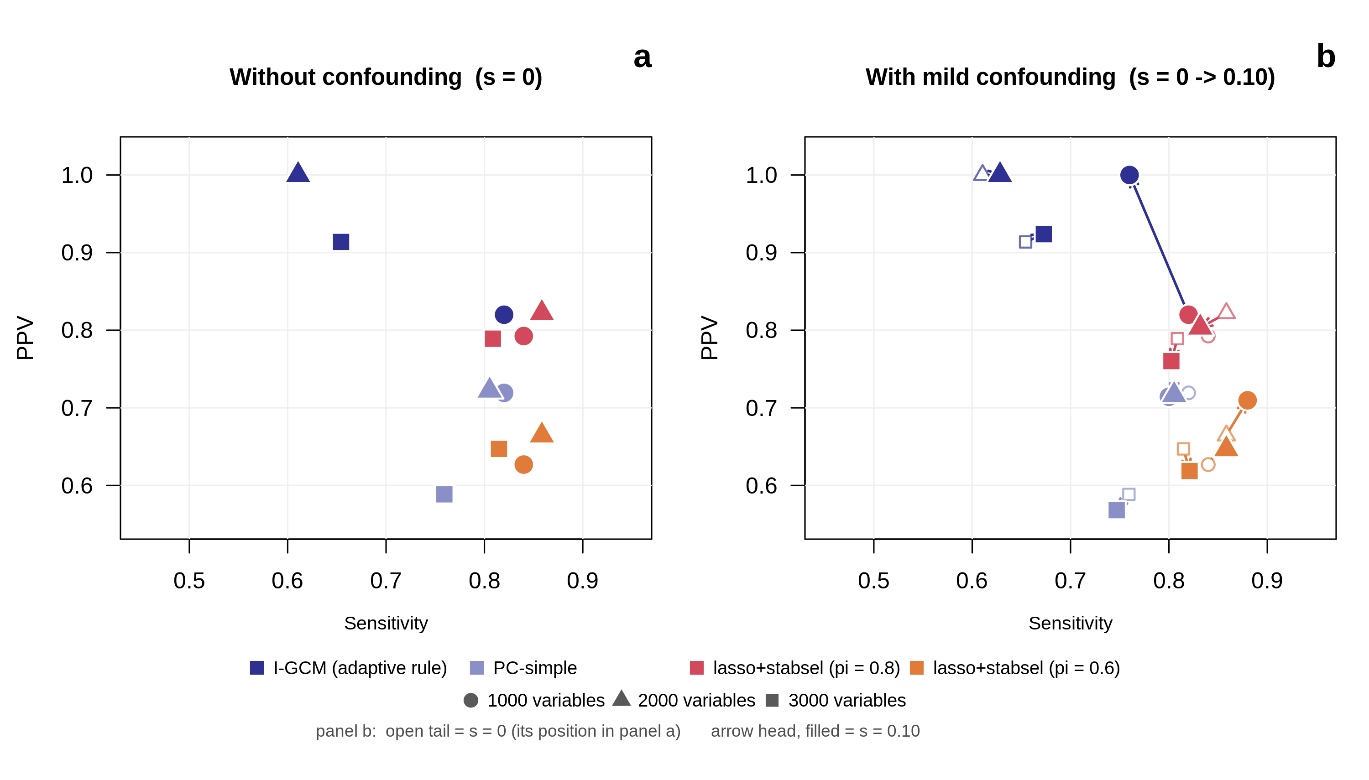

Fig. S2 Benchmark analysis against comparator causal inference and association-based methods. a: scenarios without unobserved confounding; b: scenarios with unobserved confounding
